# Enterovirus D68 epidemic, UK, 2018, was caused by subclades B3 and D1, predominantly in children and adults respectively, with both subclades exhibiting extensive genetic diversity

**DOI:** 10.1101/2021.12.09.21267508

**Authors:** Hannah C. Howson-Wells, Theocharis Tsoleridis, Izzah Zainuddin, Alexander W. Tarr, William L. Irving, Jonathan K. Ball, Louise Berry, Gemma Clark, C. Patrick McClure

**Author notes:** **Address for correspondence:** C. Patrick McClure, University of Nottingham, West Block, A Floor, Queens Medical Centre, Nottingham, NG7 2UH, UK.

## Abstract

Enterovirus D68 (EV-D68) has been recently identified in biennial epidemics coinciding with diagnoses of non-polio acute flaccid paralysis/myelitis (AFP/AFM). We investigated the prevalence, genetic relatedness and associated clinical features of EV-D68 in 194 known EV positive samples from late 2018, UK. EV-D68 was detected in 83 (58%) of the 143 EV positive samples. Sequencing and phylogenetic analysis revealed an extensive genetic diversity, split between subclades B3 (n=50) and D1 (n=33), suggesting epidemiologically unrelated infections. B3 predominated in children and younger adults, and D1 in older adults and the elderly (p=0.0009). Clinical presentation indicated causation or exacerbation of respiratory distress in 91.4% of EV-D68-positive individuals, principally: cough (75.3%), shortness of breath (56.8%), coryza (48.1%), wheeze (46.9%), supplemental oxygen required (46.9%) and fever (38.9%). Clinical features were not distinguished by subclade. Two cases of AFM were observed, one with EV-D68 detectable in the cerebrospinal fluid, otherwise neurological symptoms were rarely reported (n=4).

## Introduction

The Enterovirus (EV) genus within the non-enveloped ssRNA Picornaviridae family includes seven species infecting humans: three rhinovirus species (Rhinovirus A – Rhinovirus C) and four enterovirus species (Enterovirus A-Enterovirus D) (1). These EV species are further divided into distinct serotypes, prefixed with ‘A’, ‘B’, ‘C’ or ‘D’ to indicate their species assignment. Enterovirus infections in humans are caused by in excess of 100 serotypes and manifest as a diverse array of symptoms from asymptomatic infection to respiratory, epidermal, gastrointestinal and neurological presentations (2). Globally EV infections represent a considerable burden of morbidity and mortality (2, 3) and they remain the leading cause of viral meningitis worldwide.

Since its recent re-emergence, enterovirus D68 (EV-D68) has been associated with biennial (2014, 2016 and 2018) seasonal epidemics of respiratory infections and, more rarely, acute flaccid myelitis (AFM) across Northern America (4-6), with subsequent identification of similar patterns globally (7-9). There are limited studies looking at the true burden EV-D68. Multiple contemporary strains of EV-D68 have been associated with neuropathology in mouse models following dissemination to the central nervous system (10, 11) with others suggesting this is a universal EV-D68 phenotype including the archetypal 1962 Fermon strain (12). Despite its neurotropic nature, EV-D68 is primarily an infection of the respiratory tract and in contrast to many other enterovirus species, transmission is primarily by airborne routes and fomites (13, 14). EV-D68 is infrequently found in faeces (15) and is not thought to spread by the faecal-oral route as it is acid labile (2, 16). There are four identified EV-D68 clades, designated A-D, with subclades B3 and D1 contemporaneously predominating in global sequence data (8, 17-21). Evidence is emerging to suggest severity of EV-D68 infection is associated with the presence of other co-morbidities (22); these individuals are more likely to present with complications associated with EV-D68 infection. This large retrospective study also highlighted an upsurge in D1 cases in the adult population at a time when B3 and D1 were co-circulating.

Serological studies have retrospectively indicated an increasing prevalence of EV-D68 since at least 2006 (13, 23, 24), highlighting a growing importance and foothold in the roster of common childhood infections. The detection of enterovirus antibodies in the cerebrospinal fluid (CSF) of patients with an AFM diagnosis (25) provides further evidence to link enterovirus infection and AFM, however EV-D68 RNA is rarely detected in CSF (7, 26-28)

Enterovirus infections are now routinely diagnosed by molecular methods and are further sub-typed by neutralization or sequence based methods (2). Sequencing of the VP1 gene, encoding one of four viral capsid proteins, is considered the current diagnostic typing ‘gold-standard’ (29), rather than the highly conserved 5’ untranslated (5’ UTR) genomic region due to recombination events observed within this genomic region (30). Perhaps due to the recent re-emergence of EV-D68 after a period of apparent low prevalence [25, 26], many nucleic acid-based diagnostic assays perform relatively poorly in the detection of this subtype [27].

This study retrospectively re-screened EV positive samples from patients assessed at a regional UK hospital between September and December 2018, to determine the prevalence, genetic relatedness and clinical characteristics of EV-D68 infection. Two simple, novel EV-D68-specific RT-PCR assays were designed for both qualitative detection and sequencing of the VP1 gene. VP1 sequences were compared to the entire EV-D68 global dataset and clinical characteristics audited to assess clade phenotype.

## Methods

### Specimens and ethics

Original samples previously underwent routine diagnostic testing for EV RNA by Nottingham University Hospitals (NUH) Clinical Microbiology department, East Midlands, UK, between 2nd September and 15th December 2018. This included total nucleic acid (TNA) extraction (bioMérieux™ NucliSENS® easyMAG® system) in a 50 μl elution volume and EV RNA detection by the AusDiagnostics Respiratory Viruses (16-well) (REF 20602) and/or Viral 8-well (Ref 27093) assays for the High-Plex 24 system (REF 9150). All TNA extracts were subsequently stored at -80°C.

To prevent bias on analysis, TNA extracts were deduplicated to ensure only one specimen per patient was included with neurological panel positives preferentially retained. If multiple TNA extracts were available from the same anatomical site, the earliest TNA extract was selected. All available residual TNA extracts of sufficient volume were included here (n=194) and derived from: throat swabs (n=117) and nasopharyngeal aspirates (n=41), swabs from other sites (n=19), CSF (n=7), sputum (n=4), faeces (n=2), bronchiolar lavage (n=2), whole blood (n=1) and endotracheal secretions (n=1). Ethical approval for the use of residual material and association with anonymized patient information was locally approved under the Nottingham Health Science Biobank Research Tissue Bank, REC reference 15/NW/0685.

### RT-PCR, Primer Design and sequencing

cDNA was synthesized and amplified from TNA extracts as previously described (31). Samples were targeted with a generic 5’ UTR based assay generating a ∼480 bp product as previously described (31, 32), and two novel EV-D68 specific assays: EV-D68 assay primers were designed following alignment of all available complete EV-D68 genomes (n=348) published on Genbank by November 2018 (NCBI:txid42789). Two overlapping amplicons were generated to ensure full coverage of the EV-D68 VP1 gene: primers D68_1442F (CCCTGATAATACCTTGGATYAGTGG) combined with D68_2267R (GCTGARTTAATRCACATAAARGGTAT) and D68_2167F (GACACTNCARGCAATGTTYGT) with D68_2762R (CCTGTRTTRCAYTTRCAYCTTGCTAT) generated circa 820bp and 600bp products respectively. PCR products of expected size were diluted 1:10 in molecular grade water and subjected to Sanger sequencing (Source BioScience, UK) with primers D68_2267R and D68_2167F as appropriate. Sequence identity was assessed using NCBI Standard Nucleotide BLAST (BLASTn) and the Genome Detective online Enterovirus Genotyping Tool (Version 2.9) (33). Complete VP1 sequences were uploaded to Genbank under accession numbers MZ576283 - MZ576352.

### Phylogenetic and statistical analyses

All publicly available complete VP1 sequences were retrieved from Genbank in June 2021 and aligned with the study sequences using Geneious Prime 2019.2.3. A phylogenetic tree was constructed from the alignment in IQTREE2 (34) by the Maximum Likelihood method with the GTR+F+I model and 1000 SH-aLRT bootstraps. Clinical features were compared by genotype using a binary logistic regression model in GraphPad Prism v.9.2.0.

## Results

### Enteroviral epidemiology in Nottingham, UK, Autumn 2018

The AusDiagnostics respiratory virus panel allows for non-specific detection of both rhinovirus (RV) and enterovirus (non-differentiated dual target: RV/EV), and specific detection of EV, in TNA. Detection of RV/EV and EV in respiratory specimens increased from week 36 of 2018 (early September) and EV diagnoses remained elevated until week 45 (November, Figure 1). RV/EV numbers remained high after the decline in EV, as had been seen for the corresponding period of 2017 (data not shown). Retrospective analysis of EV positive TNA was performed on samples received for routine testing during this peak period of 2018, between weeks 36 and 50 inclusively. In total, 308 positive patients were identified, of which 194 residual TNA extracts had sufficient volume for further analysis.

**Figure 1:**
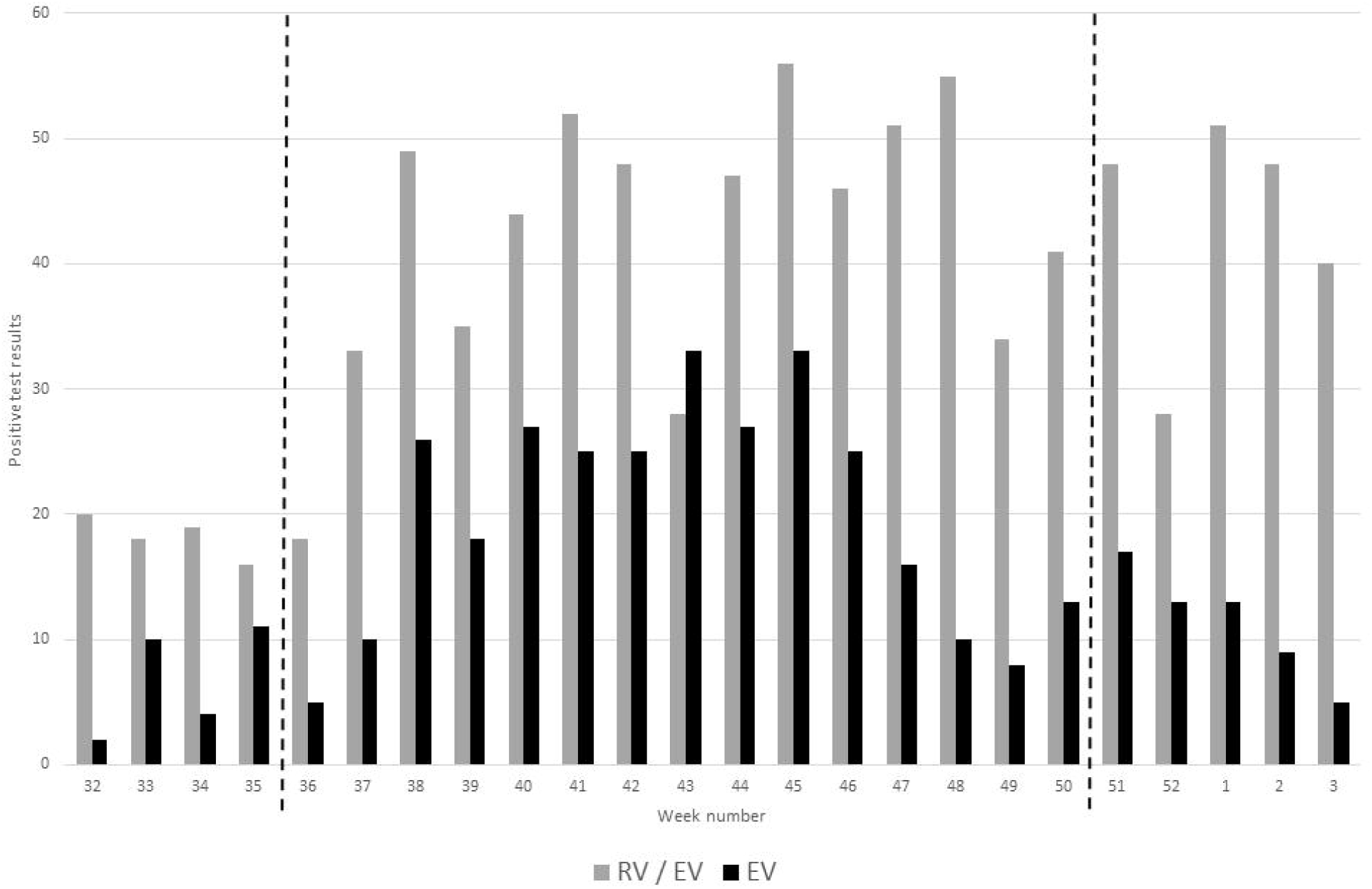
The number of respiratory specimens from which rhinovirus/enterovirus (RV/EV) and enterovirus (EV; species A-D) were detected between 2018 week 32 and 2019 week 3, as reported by NUH NHS Trust, Nottingham, UK. TNA from samples received from weeks 36 to 50 (marked by dashed lines) were subjected to further analysis.

To investigate the prevalence of EV-D68 in the cohort, two novel EV-D68 specific primer combinations, amplifying the VP1 capsid gene, were utilized. An additional novel pan-enterovirus specific control assay targeting the conserved 5’ UTR region was also employed (31).

Of these 194 EV clinical-assay-positive extracts, our study assays confirmed EV in 143 (73.7%). Of the confirmed EV positive samples, EV-D68 RNA was detected in 58.0% of cases (n=83) as determined by sequencing of products from either or both VP1 study assays.

The diagnostic results for the 51 (26.3%) TNA extracts where EV RNA was not detected by our in-house EV assays were reviewed. All had either a high ratio of combined RV/EV viral load to EV specific viral load, indicating potential cross-reactivity with rhinoviruses in the EV specific AusDiagnostics assays, three of which were confirmed by sequencing; or alternatively a low EV value on initial screening, suggesting a low EV viral load in the original sample below the lower limit of detection of the study assays or degradation of RNA within the TNA extract on repeat freeze thaw.

### Enterovirus D68 epidemiology, 2018 weeks 36 to 50

All amplicons were sequenced by Sanger and typed using the Genome Detective online tool (33). A full VP1 sequence was amplified and used for subsequent phylogenetic analysis in 70 of 83 (84.3 %) samples but sequencing of a partial VP1 region in isolation still allowed for successful subtyping. Co-circulation of two EV-D68 subclades was observed as reported concurrently elsewhere in Europe (18-20), with 50 subclade B3 and 33 subclade D1 positives identified (Figure 2). EV-D68 B3 predominated in the peak epidemic period of weeks 40 and 41, whilst strains belonging to the less prevalent subclade D1 were present throughout the study period, with peaks seen in weeks 38 and 43 (Figure 2).

**Figure 2:**
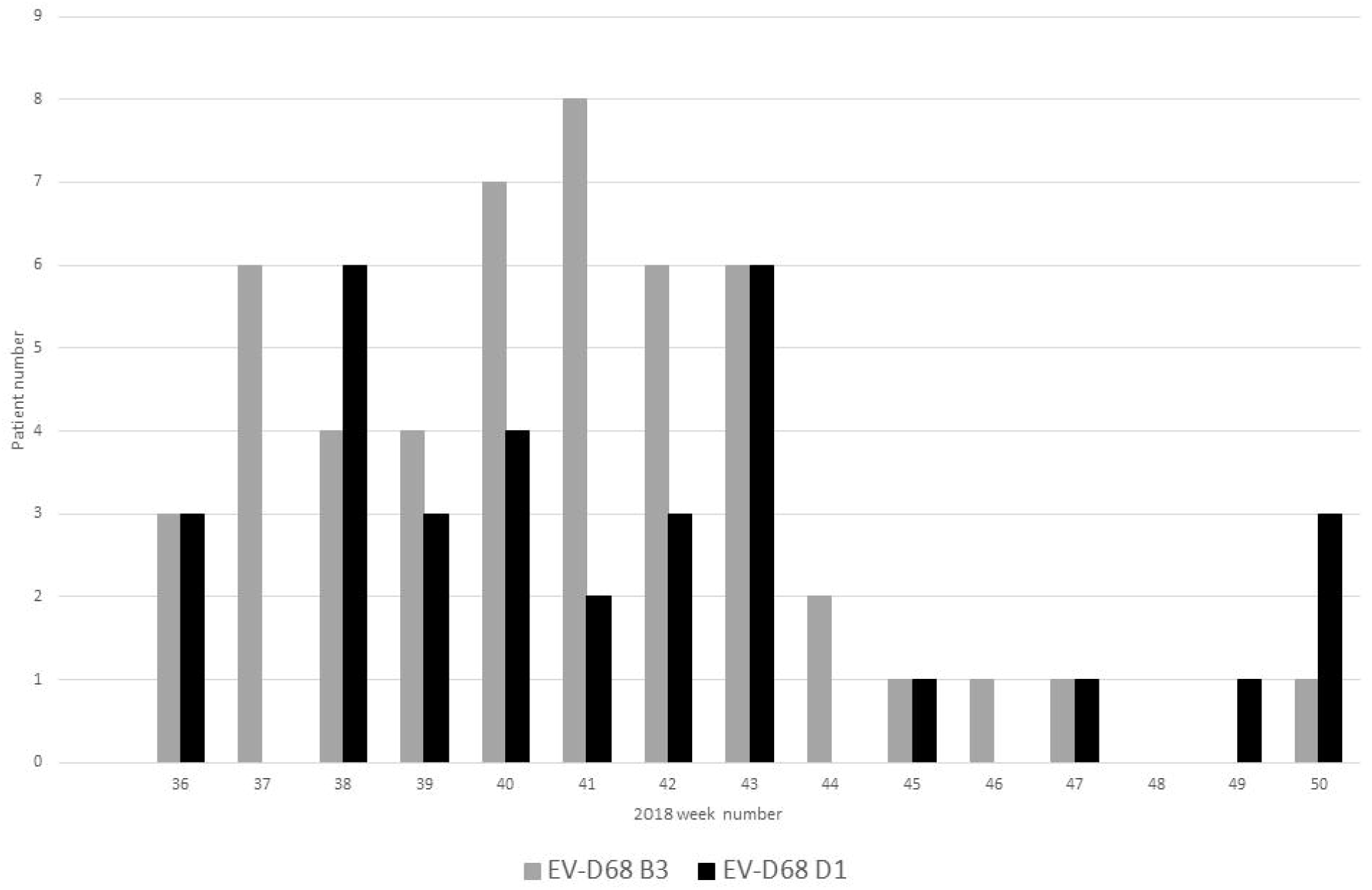
EV-D68 prevalence by individual patient and clade, from weeks 36 to 50 of 2018 at NUH NHS Trust, Nottingham UK.

Patients with EV-D68 B3 were significantly younger than those infected with subclade D1 (p value = 0.0009, Table 1) with 64.0 % (n=32) of B3 positive patients ≤18 years of ages, and % (n=25) of those patients under five years (figure 3). In contrast, 91% (n=30) of patients with EV-D68 subclade D1 were >18 years, with the majority (75.6%, n=25) of patients aged 45 years and over (Figure 3).

**Table 1.**
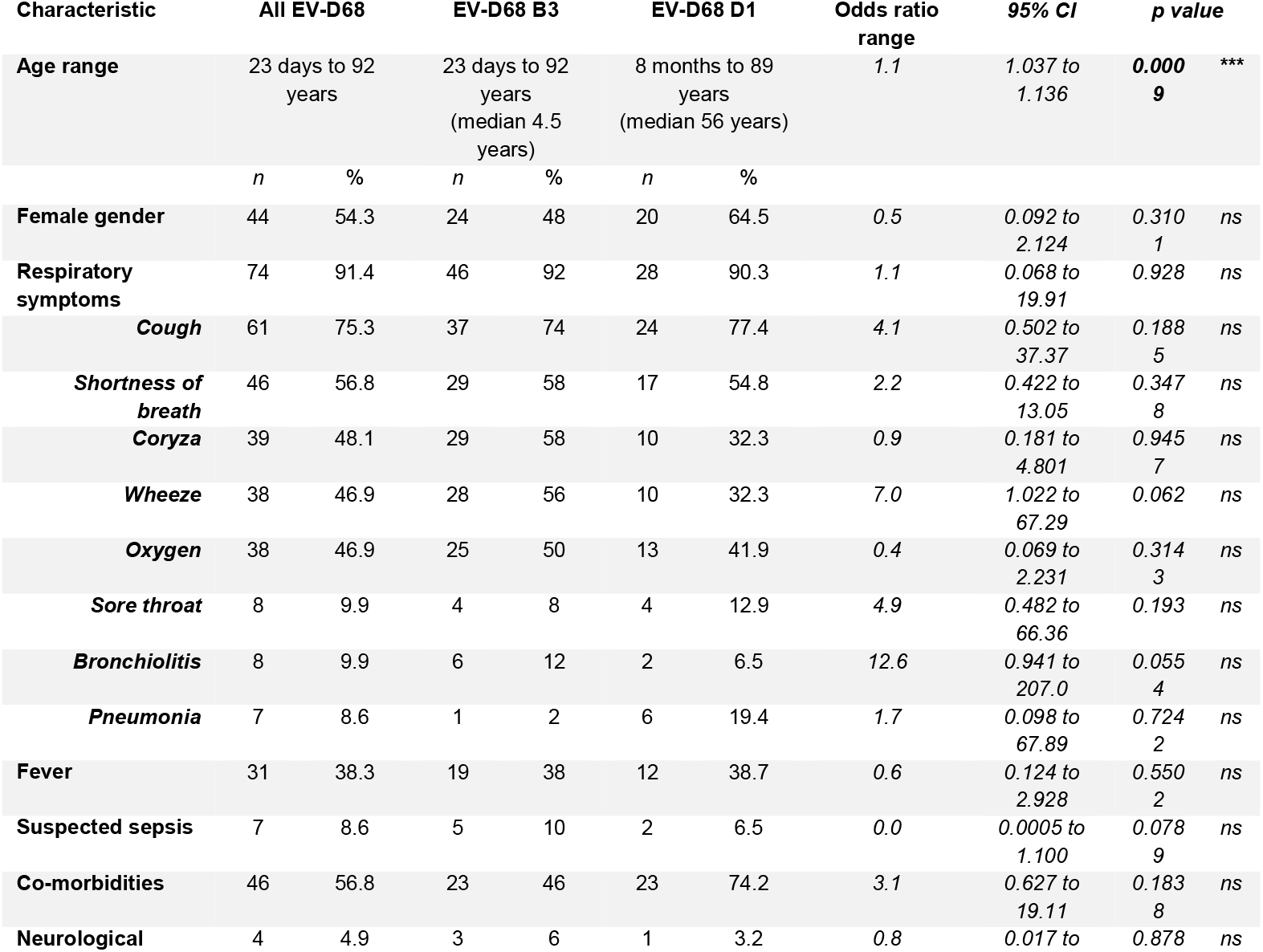
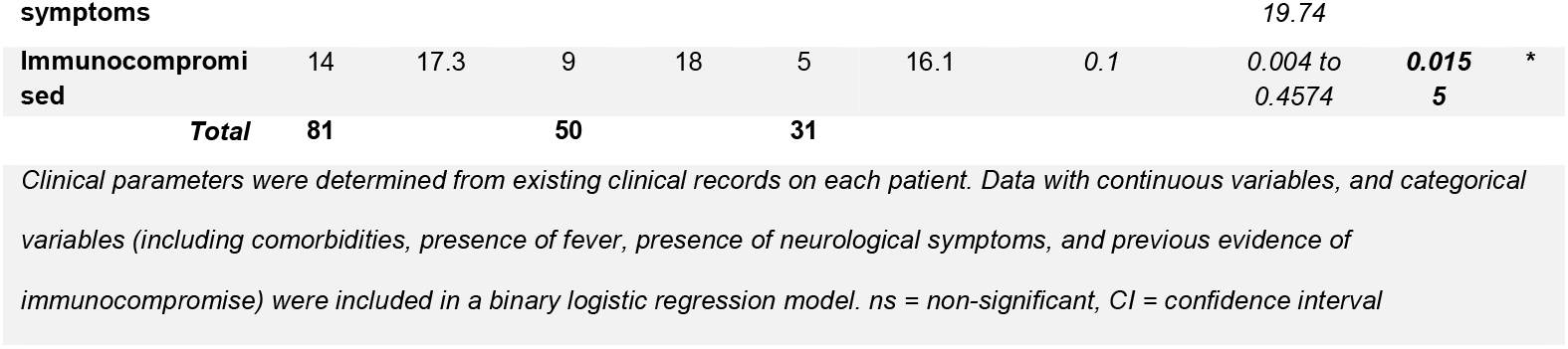
Clinical features of EV-D68 patients, Nottingham, UK, Autumn 2018

**Figure 3:**
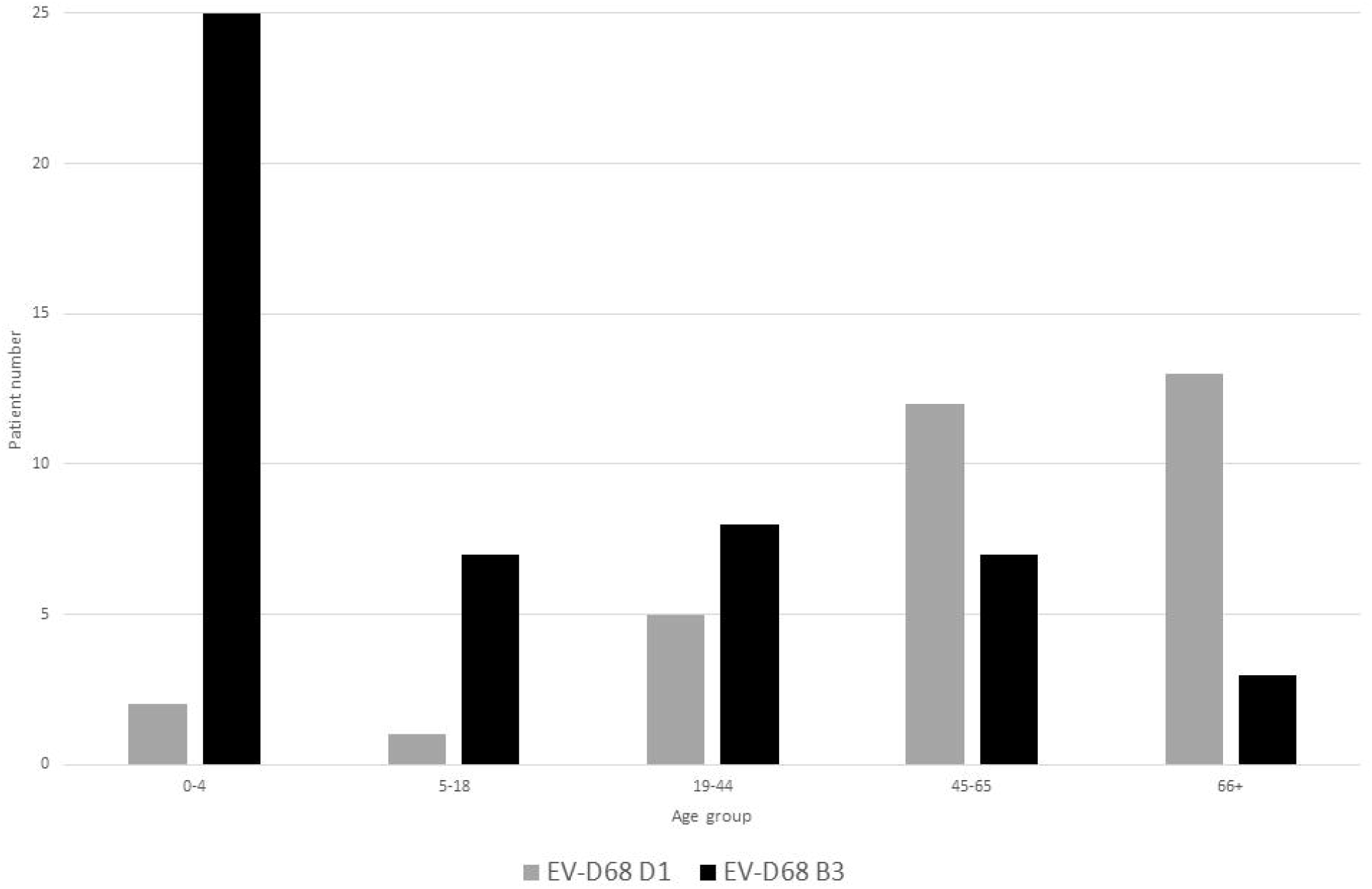
Age distribution of patients infected with EV-D68, stratified by subclade (B3 and D1).

### Clinical features of EV-D68 positive patients

Detailed clinical information was available for 81 of 83 EV-D68 positive individuals and analysis of a range of these features associated with either EV-D68 genotype D1 or genotype B3 was performed using binary logistic regression (Table 1). This regression model indicated that greater age was associated with EV-D68 D1 infection, while immunocompromise significantly associated with B3 infection. Overall, the model correctly predicted 85.19% of cases. Respiratory symptoms were present for 91.4 % of patients, most commonly a cough (75.3%), shortness of breath (56.8%), coryza (48.1%), wheeze and a requirement for oxygen (both 46.9%). Fever was also frequently reported (38.3%) as were co-morbidities (56.8%). Neurological symptoms were reported in only four cases (4.9%), including two cases of AFM, one in a young adult and the other in a 1-year old child. In both cases EV-D68 clade B3 was detected; in the CSF of the adult and, unusually, in the faeces of the paediatric AFM case. No other CSF or faecal samples in the study period were found to be EV-D68 positive. The adult AFM patient required intensive care as did five other patients – another young adult and four children aged 0-9 years. All five patients were infected with EV-D68 clade B3.

### Genetic epidemiology of Enterovirus D68 in Nottingham, UK, between September and December 2018

To investigate the genetic relationship of the study EV-D68 strains, a phylogenetic tree with our 70 complete VP1 sequences and the entire available global dataset of c.1500 sequences (Supplementary Figure 1) was constructed. Clade D1 VP1 sequences presented as a closely related group within the total dataset (Figure 4), whilst B3 sequences were split across two distinct phyletic groups (Figure 5 and 6).

**Figure 4:**
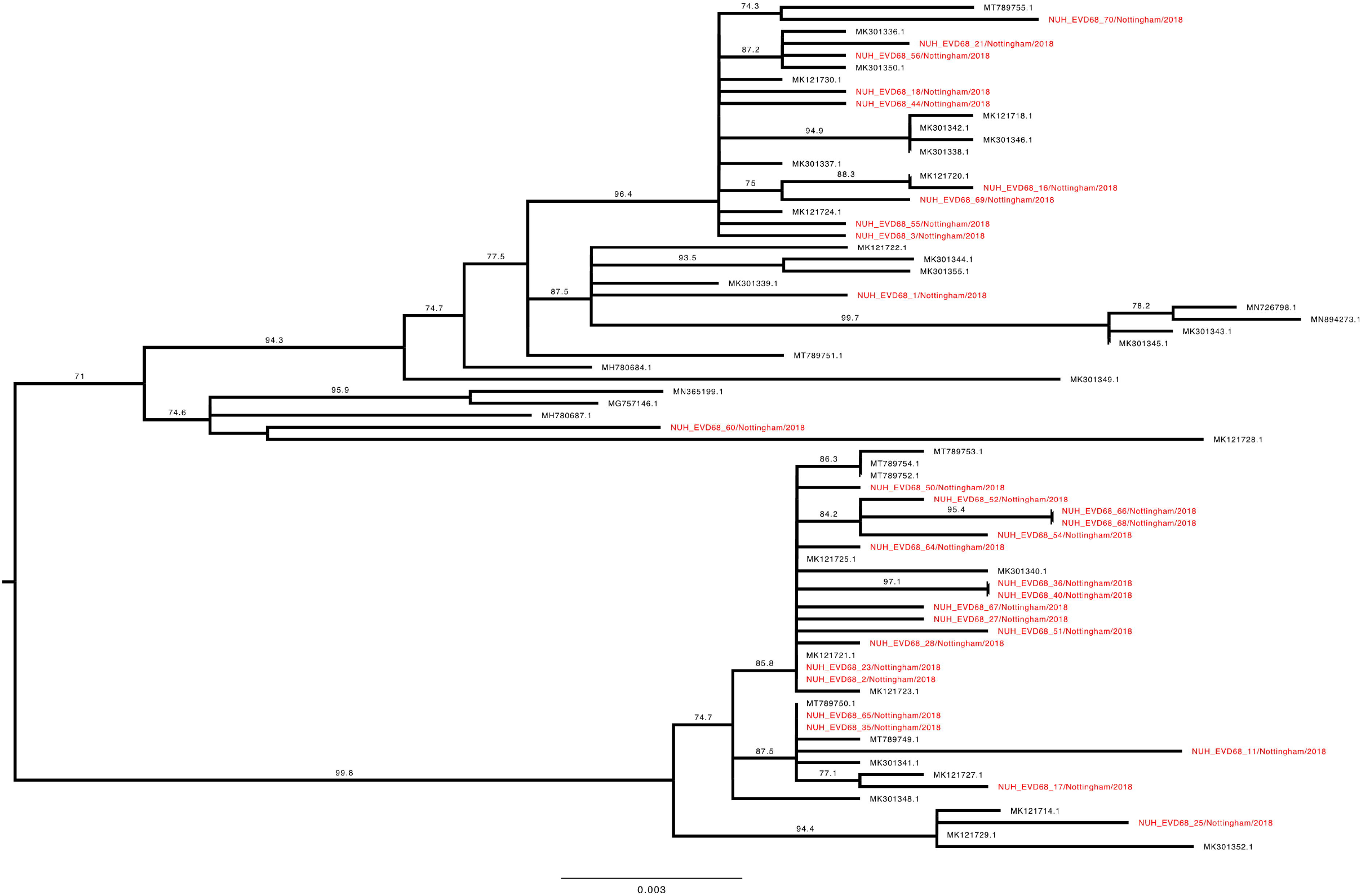
Phylogenetic relationship by maximum likelihood method of Nottingham, UK, 2018 complete EV-D68 VP1 sequences (927-930 bp) designated genotype D1 (colored in red) with all closely related publically available genomes retrieved from Genbank in June 2021 (identified by accession number). The phylogeny depicted is a subtree of a complete tree with entire study and global sequence dataset presented in the appendix (Supplementary Figure 1). Numbers above individual branches indicate SH-aLRT bootstrap support, with values less than 70 not shown. Branch lengths are drawn to a scale of nucleotide substitutions per site, with scale indicated.

**Figure 5:**
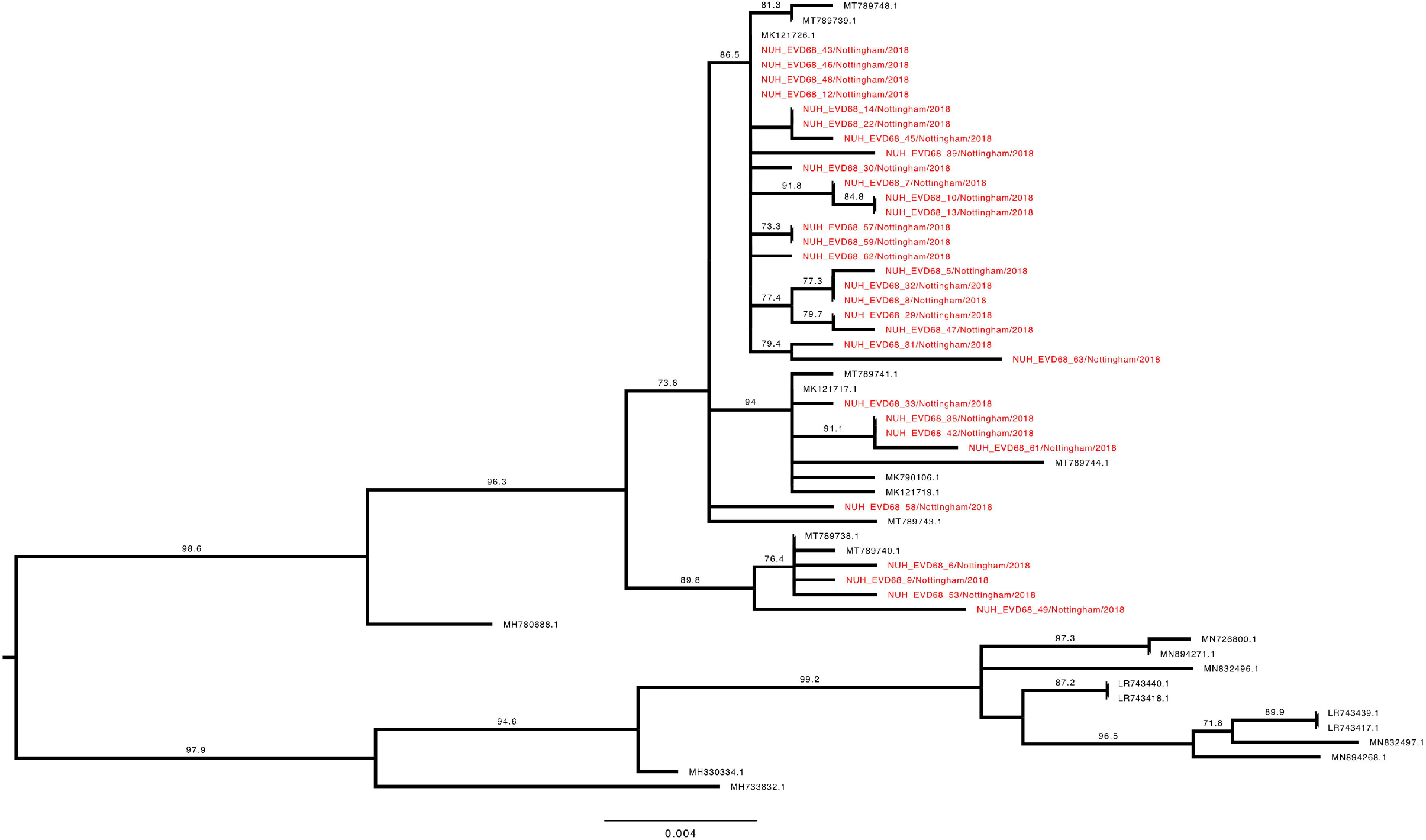
Phylogenetic relationship by maximum likelihood method of Nottingham, UK, 2018 complete EV-D68 VP1 sequences (927-930 bp) designated genotype B3, subgroup 1 (coloured in red) with all closely related publically available genomes retrieved from Genbank in June 2021 (identified by accession number). The phylogeny depicted is a subtree of a complete tree with entire study and global sequence dataset presented in the appendix (Supplementary Figure 1). Numbers above individual branches indicate SH-aLRT bootstrap support, with values less than 70 not shown. Branch lengths are drawn to a scale of nucleotide substitutions per site, with scale indicated.

**Figure 6:**
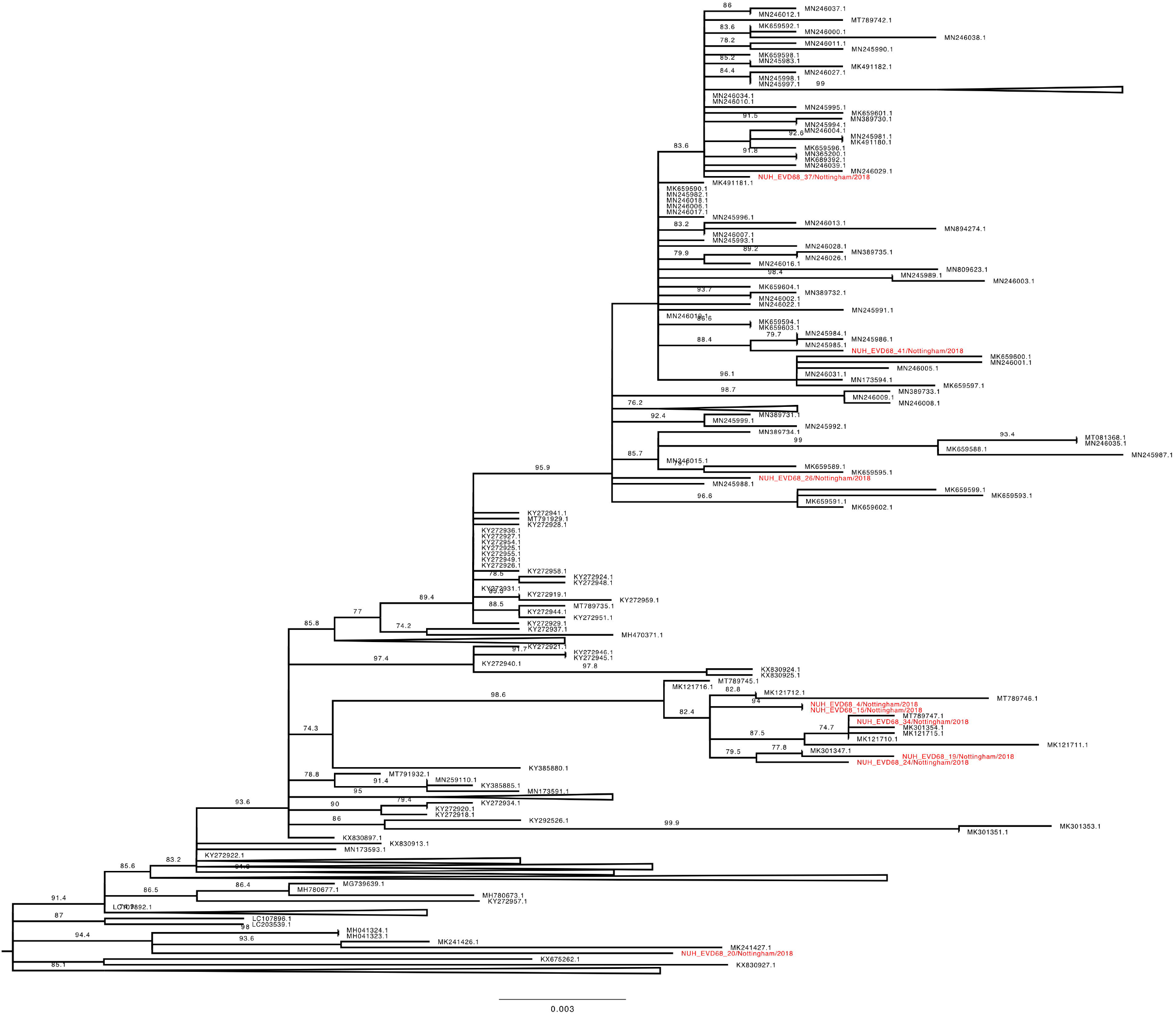
Phylogenetic relationship by maximum likelihood method of Nottingham, UK, 2018 complete EV-D68 VP1 sequences (927-930 bp) designated genotype B3, subgroup 2 (coloured in red) with all closely related publically available genomes retrieved from Genbank in June 2021 (identified by accession number). The phylogeny depicted is a subtree of a complete tree with entire study and global sequence dataset presented in the appendix (Supplementary Figure 1), with well supported branches containing no study sequences collapsed to improve clarity. Numbers above individual branches indicate SH-aLRT bootstrap support, with values less than 70 not shown. Branch lengths are drawn to a scale of nucleotide substitutions per site, with scale indicated.

The VP1 subtree harbouring all the D1 sequences from this study (Figure 4) was most closely out-grouped, yet distinct from, both prior-2016 isolates from East Asia (e.g. MH780679 and KY358058), a contemporary 2018 French sample (MK121713 (19)) and later-2019 isolates from Europe (e.g. LR743441 (17)). The subtree containing all 30 full-length VP1 D1 sequences from this study and 39 database references could be subdivided into two further phyletic groups. The first (Figure 4, lower half) contained exclusively contemporary European isolates from investigations in France and Italy ((18, 19), whereas the other (Figure 4, upper half) additionally contained a well-supported subgroup with four 2017 sequences from the USA and Hong Kong, a 2018 French isolate (MK121728 (19)) and a single Nottingham sequence. Interestingly, this latter patient (NUH_EVD68_60) was the only one in the cohort to report recent travel history to East Asia. Our study sequences frequently shared a closer genetic relationship with contemporary isolates from continental Europe than with each other. For example, samples NUH_EVD68_21, 23 & 25 were all sampled within 24 hours of each other, yet appeared in discrete, well-supported regions of the tree with a closer genetic identity to various European isolates than each other (Figure 4).

The Nottingham EV-D68 B3 sequences (Figures 5 and 6) presented a similar picture of co-mingling with contemporary European isolates, with for example NUH_EVD68_34 residing in a well-supported cluster of EV-D68 B3 sequences with three French and one Italian strain (Figure 6). Whereas the chronologically adjacent UK sequence NUH_EVD68_33 sampled on the previous day, appeared in an entirely different but equally well-supported sub-clade of B3 (Figure 5).

Recent travel history was also noted for other patients in both clades (NUH_EVD68_4, 7, 20 & 27, Figures 4-6). Of note, NUH_EVD68_20 (Figure 6), with recent Spanish travel history, presented in a distinct supported phyletic group with four Chinese sequences from 2016 and 2017 (35). The other cases, with travel histories to the USA, Denmark and Spain did not present any distinctive sub-groupings with reference sequences. However, NUH_EVD68_4 (Figure 6), also with recent Spanish travel history, presented an identical VP1 sequence to NUH_EVD68_15 received 13 days later. Both these patients were 5 years old and shared the same hometown and outer UK postcode. There were eight other instances of identical pairs or clusters of study sequences, but clinical details did not suggest a direct familial or geographical association. Most were relatively closely sampled in time, but NUH_EVD68_35 and 65 were received two months apart in different but adjacent UK outer postcodes.

Taken together, these observations would suggest both Europe-wide and local circulation of multiple strains of both B3 and D1 clades underlying the Autumn 2018 EV-D68 epidemic.

## Discussion

Enteroviruses can cause a wide range of severe clinical manifestations, notably in young children and are a leading cause of meningitis globally (2). Continued surveillance of enteroviruses within the UK despite the elimination of Polio is still highly relevant, notably highlighted by the recent resurgence and recurrent epidemic seasonality of EV-D68 with associated AFM globally and nationally in the UK (36-39).

Growing recognition of AFM and enteroviral association in Autumn 2018 prompted us to further investigate enterovirus positive samples using novel in house RT-PCR assays and Sanger sequencing. Whilst this approach is well established and more recently over-shadowed by whole genome sequencing using deep sequencing methods, it provides a flexible, inexpensive and fast route to enteroviral typing and epidemiology in a clinical setting.

We found a significant burden of EV-D68 infection in our 2018 study period between September and December, coinciding with an overall increase in EV/RV positivity, responsible for 58.0% of 143 confirmed enteroviral positives, over ten-fold higher than its overall recent EU prevalence of 4.53% (2). It is difficult to determine the context of this spike in EV-D68 positivity locally, relative to both the rest of 2018 and other years, without routine typing of all enterovirus positive respiratory samples in addition to the more frequently assessed neurological symptom-associated enteroviral infections (2, 26, 31, 40). EV-D68 is an endemic infection maintained in the population and is still detected in Europe out of the currently expected biennial peak years (17), but resources were not available to routinely type respiratory samples outside of our focused study period.

We targeted the highly variable and type-indicative enteroviral VP1 region (26) with novel primers specific to EV-D68 to not only discriminate EV-D68 infection (no cross-reactivity was observed with other enteroviruses), but also determine the relative contribution by circulating clades. To our knowledge, this allows us to present the largest fully typed EV-D68 cohort from a single centre and epidemic season. Whilst both clades were present throughout our sampling period, an approximately 60:40 predominance of clade B3 relative to D1 was observed. This was the reverse of clade prevalence seen in other European studies of the 2018 EV-D68 season in France (19) and Italy (18), but in agreement with other investigations of previous seasons in France (20), Italy (21) and more generally in Europe post-2018 (17).

Universal typing and relatively large cohort size enabled us to investigate EV-D68 clade association with clinical characteristics with more statistical power than most other studies to date. Here we found a highly significant association between age and clade type, with clade B3 predominantly affecting children and adolescents, whilst conversely D1 was seen in older adults and the elderly. This finding is in agreement with previous observations (8, 19, 21), but only one other study of sufficient size and scope to assign statistical significance (41).

EV-D68 seroprevalence in adults is almost universal with primary infection seen frequently in the first two years of life (23, 24). The predominance of the novel D1 clade in hospitalized older adults may indicate an evolutionary adaptation of EV-D68 to evade a pre-existing immune response and facilitate repeat infection. Recent and rapid evolution of antigenic regions of the outer viral capsid corresponding to neutralizing epitopes has been suggested to be implicated (8, 21, 41). Lineage specific protection determined by a primary infection influencing virus-type age distribution has been described for several other viral infections, notably Influenza A and B (42, 43).

Despite the significant differences in age profile for each clade and our cohort size, no difference was seen in clinical presentations recorded, in agreement with others (20). The general clinical picture of symptoms indicative of both upper and lower respiratory tract infections was consistent with other studies (18, 19, 21, 28, 44). Although all severe cases, including those with neurological involvement such as AFM, presented with clade B3, their infrequency prevents robust analysis and all were in younger individuals shown to be more likely infected by clade B3. Indeed AFM is not restricted to the B3 clade (4, 45, 46) and is perhaps more likely associated with the predominant local strain infecting a naïve population and overall infection rates, rather than recently acquired properties as hypothesized by some (46) and thus not a recently acquired phenotype (12).

Perhaps more notable in both of the AFM cases was the infrequently reported viral genome detection in CSF and faeces, with EV-D68 being a predominantly respiratory pathogen and thus typically detected by sampling the respiratory tract (2, 47). This finding supports a role for viral replication disseminated from the respiratory tract in EV-D68-associated AFM, although EV-D68 detection in sewage in the UK (48) and elsewhere (49) suggests faecal shedding may not be an unusual feature of infection in general.

We observed a considerable genetic diversity within the VP1 regions of both the B3 and D1 clades within our single epidemic season analysis, consistent and co-mingling with other contemporary European cohorts (18, 19). In conjunction with the post-summer spike in case incidence, this is suggestive of a high level of community infection seeded by many regional and international introductions as described in more detail elsewhere (41) and foreshadowing SARS-CoV-2 pandemic dynamics (50). Our complete VP1 sequencing in conjunction with a detailed clinical audit provides greater granularity than many other studies. Notably an individual with immediate East Asian travel history presented an EV-D68 VP1 sequence distinct from any other European isolates recorded to date, suggesting direct intercontinental importation. Other cases with recent summer travel history in Europe, combined with well-supported genetically related sequences from both Europe and the immediate local neighbourhood, are anecdotally suggestive of acquisition on holiday and onward transmission thereafter. Another case with Spanish travel history clustered exclusively with sequences from previous years in China, suggestive of considerable yet undetected intercontinental and international transmission chains occurring.

However, with minimal global surveillance in comparison to that seen with SARS-CoV-2, assessing onward transmission from such introductions is difficult to definitely ascertain in the short term, with prevailing introductions apparent only in subsequent epidemic periods. Yet despite this lack of resolution, EV-D68 VP1 sequencing assured local infection control teams in retrospect that individuals sharing hospital wards did not harbour genetically identical viruses and thus nosocomial transmission was unlikely and infection prevention measures in place were effective.

We acknowledge several limitations in the work presented, principally the relatively short single time-period assessed. Continuous and routine typing of all candidate EV positive specimens received from an earlier outset would have provided a more complete picture and understanding of the evolving local EV-D68 burden. We thus could not assign a start and end point to our EV-D68 epidemic season. Recent initiatives such as the European non-polio enterovirus network (26, 36, 40) to which NUH NHS Trust Clinical Microbiology have contributed, will undoubtedly assist in increasing such surveillance. Working retrospectively with available diagnostic surplus further reduced both the sampling in the selected time period and also generally meant only single patient samples were available, thus for example general faecal shedding from patients with respiratory symptoms could not be investigated. We also may have overlooked some low titre EV-D68 positive patients by not re-screening routine diagnostic assay negative samples. Sub-optimal performance in diagnosing EV-D68 and other EV by our chosen routine diagnostic assay and others has indeed been observed (40).

In summary, EV-D68 contributed significantly to the burden of enteroviral respiratory disease treated at our regional UK hospital between September and December 2018. Underlying this period of high prevalence were genetically distinct strains of clade B3 and D1 EV-D68, which differed in their infected host’s age group, but not in clinical presentation. Despite considerable contemporary concern and focus on the severe associated outcomes of EV-D68 infection, ultimately confirmed AFM cases were rare even in this largely hospitalized cohort. Nevertheless, the apparently increasing burden of EV-D68 and diverse, rapidly evolving genome demands continued heightened surveillance.

## Data Availability

All data produced in the present work are contained in the manuscript and sequences generated deposited in GenBank under accession numbers MZ576283 - MZ576352. Additional data produced in the present study are available upon reasonable request to the authors.

## Acknowledgments

None to report

## Disclaimers

None to report

## Figure Legends

**Supplementary Figure 1.**
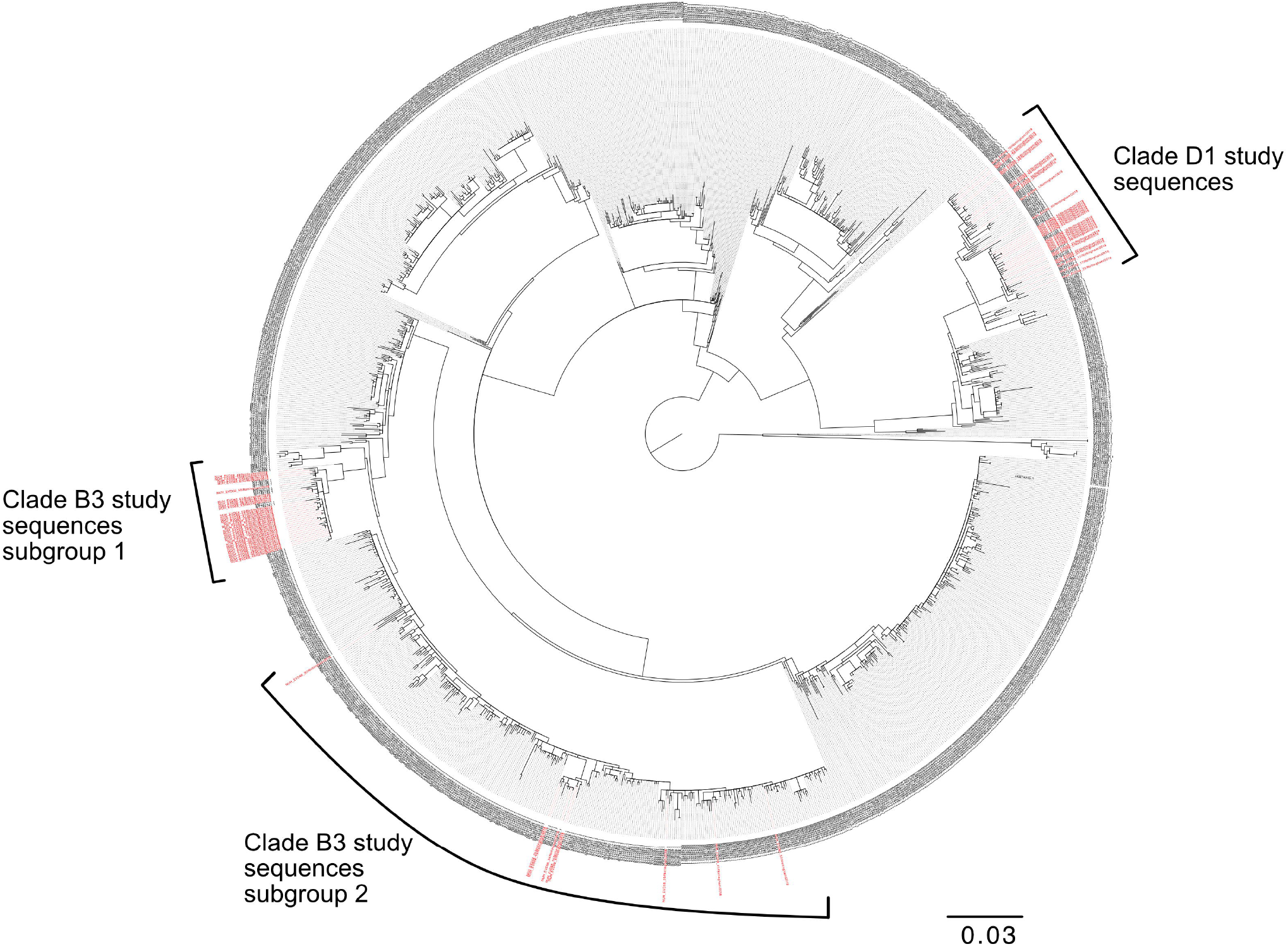
Phylogenetic relationship by maximum likelihood method of Nottingham, UK, 2018 complete EV-D68 VP1 sequences (927-930 bp, coloured in red) with all publically available genomes retrieved from Genbank in June 2021 (identified by accession number). Position of subtrees presented as figures 4 - 6 in main text is annotated. SH-aLRT bootstrap support values have been omitted for clarity. Branch lengths are drawn to a scale of nucleotide substitutions per site, with scale indicated.

## References

1. Simmonds P, Gorbalenya AE, Harvala H, Hovi T, Knowles NJ, Lindberg AM, et al. Recommendations for the nomenclature of enteroviruses and rhinoviruses. Arch Virol. 2020 Mar;165(3):793–7.

2. Bubba L, Broberg EK, Jasir A, Simmonds P, Harvala H, Enterovirus study c. Circulation of non-polio enteroviruses in 24 EU and EEA countries between 2015 and 2017. a retrospective surveillance study. Lancet Infect Dis. 2019 Dec 20.

3. Pons-Salort M, Grassly NC. Serotype-specific immunity explains the incidence of diseases caused by human enteroviruses. Science. 2018 Aug 24;361(6404):800–3.

4. Greninger AL, Naccache SN, Messacar K, Clayton A, Yu G, Somasekar S, et al. A novel outbreak enterovirus D68 strain associated with acute flaccid myelitis cases in the USA (2012-14): a retrospective cohort study. Lancet Infect Dis. 2015 Jun;15(6):671–82.

5. Eshaghi A, Duvvuri VR, Isabel S, Banh P, Li A, Peci A, et al. Global Distribution and Evolutionary History of Enterovirus D68, with Emphasis on the 2014 Outbreak in Ontario, Canada. Front Microbiol. 2017;8:257.

6. Messacar K, Schreiner TL, Maloney JA, Wallace A, Ludke J, Oberste MS, et al. A cluster of acute flaccid paralysis and cranial nerve dysfunction temporally associated with an outbreak of enterovirus D68 in children in Colorado, USA. Lancet. 2015 Apr 25;385(9978):1662–71.

7. Kirolos A, Mark K, Shetty J, Chinchankar N, McDougall C, Eunson P, et al. Outcome of paediatric acute flaccid myelitis associated with enterovirus D68: a case series. Dev Med Child Neurol. 2019 Mar;61(3):376–80.

8. Lau SK, Yip CC, Zhao PS, Chow WN, To KK, Wu AK, et al. Enterovirus D68 Infections Associated with Severe Respiratory Illness in Elderly Patients and Emergence of a Novel Clade in Hong Kong. Sci Rep. 2016 Apr 28;6:25147.

9. Gong YN, Yang SL, Shih SR, Huang YC, Chang PY, Huang CG, et al. Molecular evolution and the global reemergence of enterovirus D68 by genome-wide analysis. Medicine (Baltimore). 2016 Aug;95(31):e4416.

10. Hixon AM, Yu G, Leser JS, Yagi S, Clarke P, Chiu CY, et al. A mouse model of paralytic myelitis caused by enterovirus D68. PLoS Pathog. 2017 Feb;13(2):e1006199.

11. Brown DM, Hixon AM, Oldfield LM, Zhang Y, Novotny M, Wang W, et al. Contemporary Circulating Enterovirus D68 Strains Have Acquired the Capacity for Viral Entry and Replication in Human Neuronal Cells. mBio. 2018 Oct 16;9(5).

12. Rosenfeld AB, Warren AL, Racaniello VR. Neurotropism of Enterovirus D68 Isolates Is Independent of Sialic Acid and Is Not a Recently Acquired Phenotype. mBio. 2019 Oct 22;10(5).

13. Vogt MR, Crowe JE, Jr. Current Understanding of Humoral Immunity to Enterovirus D68. J Pediatric Infect Dis Soc. 2018 Dec 26;7(Suppl_2):S49–S53.

14. Oberste MS, Maher K, Schnurr D, Flemister MR, Lovchik JC, Peters H, et al. Enterovirus 68 is associated with respiratory illness and shares biological features with both the enteroviruses and the rhinoviruses. J Gen Virol. 2004 Sep;85(Pt 9):2577–84.

15. Levy A, Roberts J, Lang J, Tempone S, Kesson A, Dofai A, et al. Enterovirus D68 disease and molecular epidemiology in Australia. J Clin Virol. 2015 Aug;69:117–21.

16. Foster CB, Friedman N, Carl J, Piedimonte G. Enterovirus D68: a clinically important respiratory enterovirus. Cleve Clin J Med. 2015 Jan;82(1):26–31.

17. Midgley SE, Benschop K, Dyrdak R, Mirand A, Bailly JL, Bierbaum S, et al. Co-circulation of multiple enterovirus D68 subclades, including a novel B3 cluster, across Europe in a season of expected low prevalence, 2019/20. Euro Surveill. 2020 Jan;25(2).

18. Pellegrinelli L, Giardina F, Lunghi G, Uceda Renteria SC, Greco L, Fratini A, et al. Emergence of divergent enterovirus (EV) D68 sub-clade D1 strains, northern Italy, September to October 2018. Euro Surveill. 2019 Feb;24(7).

19. Bal A, Sabatier M, Wirth T, Coste-Burel M, Lazrek M, Stefic K, et al. Emergence of enterovirus D68 clade D1, France, August to November 2018. Euro Surveill. 2019 Jan;24(3).

20. Kramer R, Sabatier M, Wirth T, Pichon M, Lina B, Schuffenecker I, et al. Molecular diversity and biennial circulation of enterovirus D68: a systematic screening study in Lyon, France, 2010 to 2016. Euro Surveill. 2018 Sep;23(37).

21. Piralla A, Principi N, Ruggiero L, Girello A, Giardina F, De Sando E, et al. Enterovirus-D68 (EV-D68) in pediatric patients with respiratory infection: The circulation of a new B3 clade in Italy. J Clin Virol. 2018 Feb -Mar;99-100:91–6.

22. Duval M, Mirand A, Lesens O, Bay JO, Caillaud D, Gallot D, et al. Retrospective Study of the Upsurge of Enterovirus D68 Clade D1 among Adults (2014-2018). Viruses. 2021 Aug 13;13(8).

23. Kamau E, Harvala H, Blomqvist S, Nguyen D, Horby P, Pebody R, et al. Increase in Enterovirus D68 Infections in Young Children, United Kingdom, 2006-2016. Emerg Infect Dis. 2019 Jun;25(6):1200–3.

24. Harrison CJ, Weldon WC, Pahud BA, Jackson MA, Oberste MS, Selvarangan R. Neutralizing Antibody against Enterovirus D68 in Children and Adults before 2014 Outbreak, Kansas City, Missouri, USA(1). Emerg Infect Dis. 2019 Mar;25(3):585–8.

25. Mishra N, Ng TFF, Marine RL, Jain K, Ng J, Thakkar R, et al. Antibodies to Enteroviruses in Cerebrospinal Fluid of Patients with Acute Flaccid Myelitis. mBio. 2019 Aug 13;10(4).

26. Harvala H, Broberg E, Benschop K, Berginc N, Ladhani S, Susi P, et al. Recommendations for enterovirus diagnostics and characterisation within and beyond Europe. J Clin Virol. 2018 Apr;101:11–7.

27. Bowers JR, Valentine M, Harrison V, Fofanov VY, Gillece J, Delisle J, et al. Genomic Analyses of Acute Flaccid Myelitis Cases among a Cluster in Arizona Provide Further Evidence of Enterovirus D68 Role. mBio. 2019 Jan 22;10(1).

28. Wang H, Diaz A, Moyer K, Mele-Casas M, Ara-Montojo MF, Torrus I, et al. Molecular and Clinical Comparison of Enterovirus D68 Outbreaks among Hospitalized Children, Ohio, USA, 2014 and 2018. Emerg Infect Dis. 2019 Nov;25(11):2055–63.

29. Nix WA, Oberste MS, Pallansch MA. Sensitive, seminested PCR amplification of VP1 sequences for direct identification of all enterovirus serotypes from original clinical specimens. J Clin Microbiol. 2006 Aug;44(8):2698–704.

30. Simmonds P, Welch J. Frequency and dynamics of recombination within different species of human enteroviruses. J Virol. 2006 Jan;80(1):483–93.

31. Howson-Wells HC, Winckles S, Aliker C, Tarr AW, Irving WL, Clark G, et al. Enterovirus subtyping in a routine UK laboratory setting between 2013 and 2017. J Clin Virol. 2020 Nov;132:104646.

32. Dierssen U, Rehren F, Henke-Gendo C, Harste G, Heim A. Rapid routine detection of enterovirus RNA in cerebrospinal fluid by a one-step real-time RT-PCR assay. J Clin Virol. 2008 May;42(1):58–64.

33. Kroneman A, Vennema H, Deforche K vd, Avoort H, Penaranda S, Oberste MS, et al. An automated genotyping tool for enteroviruses and noroviruses. J Clin Virol. 2011 Jun;51(2):121–5.

34. Minh BQ, Schmidt HA, Chernomor O, Schrempf D, Woodhams MD, von Haeseler A, et al. IQ-TREE 2: New Models and Efficient Methods for Phylogenetic Inference in the Genomic Era. Mol Biol Evol. 2020 May 1;37(5):1530–4.

35. Tang SH, Yuan Y, Xie ZH, Chen MJ, Fan XD, Guo YH, et al. Enterovirus D68 in hospitalized children with respiratory symptoms in Guangdong from 2014 to 2018: Molecular epidemiology and clinical characteristics. J Clin Virol. 2021 Aug;141:104880.

36. Fischer TK, Simmonds P, Harvala H. The importance of enterovirus surveillance in a post-polio world. Lancet Infect Dis. 2021 Jul 12.

37. Carrion Martin AI, Pebody RG, Danis K, Ellis J, Niazi S, S Del, et al. The emergence of enterovirus D68 in England in autumn 2014 and the necessity for reinforcing enterovirus respiratory screening. Epidemiol Infect. 2017 Jul;145(9):1855–64.

38. Park SW, Pons-Salort M, Messacar K, Cook C, Meyers L, Farrar J, et al. Epidemiological dynamics of enterovirus D68 in the United States and implications for acute flaccid myelitis. Sci Transl Med. 2021 Mar 10;13(584).

39. he United Kingdom Acute Flaccid Paralysis Afp Task F. An increase in reports of acute flaccid paralysis (AFP) in the United Kingdom, 1 January 2018-21 January 2019: early findings. Euro Surveill. 2019 Feb;24(6).

40. Hayes A, Nguyen D, Andersson M, Anton A, Bailly JL, Beard S, et al. A European multicentre evaluation of detection and typing methods for human enteroviruses and parechoviruses using RNA transcripts. J Med Virol. 2019 Dec 28.

41. Hodcroft EB, Dyrdak R, Andrés C, Egli A, Reist J, García Martínez de Artola D, et al. Evolution, geographic spreading, and demographic distribution of Enterovirus D68. bioRxiv. 2020:2020.01.10.901553.

42. Vieira MC, Donato CM, Arevalo P, Rimmelzwaan GF, Wood T, Lopez L, et al. Lineage-specific protection and immune imprinting shape the age distributions of influenza B cases. Nat Commun. 2021 Jul 14;12(1):4313.

43. Gostic KM, Ambrose M, Worobey M, Lloyd-Smith JO. Potent protection against H5N1 and H7N9 influenza via childhood hemagglutinin imprinting. Science. 2016 Nov 11;354(6313):722–6.

44. Schuster JE, Selvarangan R, Hassan F, Briggs KB, Hays L, Miller JO, et al. Clinical Course of Enterovirus D68 in Hospitalized Children. Pediatr Infect Dis J. 2017 Mar;36(3):290–5.

45. Kaida A, Iritani N, Yamamoto SP, Kanbayashi D, Hirai Y, Togawa M, et al. Distinct genetic clades of enterovirus D68 detected in 2010, 2013, and 2015 in Osaka City, Japan. PLoS One. 2017;12(9):e0184335.

46. Zhang Y, Cao J, Zhang S, Lee AJ, Sun G, Larsen CN, et al. Genetic changes found in a distinct clade of Enterovirus D68 associated with paralysis during the 2014 outbreak. Virus Evol. 2016 Jan;2(1):vew015.

47. Chong PF, Kira R, Mori H, Okumura A, Torisu H, Yasumoto S, et al. Clinical Features of Acute Flaccid Myelitis Temporally Associated With an Enterovirus D68 Outbreak: Results of a Nationwide Survey of Acute Flaccid Paralysis in Japan, August-December 2015. Clin Infect Dis. 2018 Feb 10;66(5):653–64.

48. Majumdar M, Martin J. Detection by Direct Next Generation Sequencing Analysis of Emerging Enterovirus D68 and C109 Strains in an Environmental Sample From Scotland. Front Microbiol. 2018;9:1956.

49. Weil M, Mandelboim M, Mendelson E, Manor Y, Shulman L, Ram D, et al. Human enterovirus D68 in clinical and sewage samples in Israel. J Clin Virol. 2017 Jan;86:52–5.

50. du Plessis L, McCrone JT, Zarebski AE, Hill V, Ruis C, Gutierrez B, et al. Establishment and lineage dynamics of the SARS-CoV-2 epidemic in the UK. Science. 2021 Feb 12;371(6530):708–12.

